# Clinic audit on Chronic Obstructive Pulmonary Disease (COPD) management in public primary care setting: Hong Kong experience

**DOI:** 10.1101/2020.11.26.20239541

**Authors:** Chen Xiao Rui Catherine, Fu Sau Nga, Leung Wing Kit, Ng Sze Wing Catherine, Kwan Wing Yan Wendy, Wong Tseng Kwong, Chan Pang Fai, Wong Man Ying Michelle, Ko Wai Kit Welchie, Liang Jun, Hui Ming Tung Eric, Li Yim Chu, Luk Wan, Chao VK David

**Affiliations:** Dept. of Family Medicine and General Out Patient Clinics, Kowloon Central Cluster, Hospital Authority, Hong Kong; Dept. of Family Medicine and Primary Health Care, Kowloon West Cluster, Hospital Authority, Hong Kong; Dept. of Family Medicine, New Territories East Cluster, Hospital Authority, Hong Kong; Dept. of Family Medicine and Primary Health Care, Hong Kong West Cluster, Hospital Authority, Hong Kong; Dept. of Family Medicine and Primary Health Care, Hong Kong East Cluster, Hospital Authority, Hong Kong; Dept. of Family Medicine and Primary Health Care, Kowloon East Cluster, Hospital Authority, Hong Kong; Dept. of Family Medicine and Primary Health Care, New Territories West Cluster, Hospital Authority, Hong Kong

**Keywords:** COPD, clinical audit, primary care, quality improvement

## Abstract

**Background:** Chronic obstructive pulmonary disease (COPD) is a common condition encountered in primary care and presents a substantial burden to the health care system. This study aimed to audit COPD care at all public primary care clinics of Hong Kong and to work out improvement strategies.

**Method:** The computer record of COPD patients aged 40 or above and had been followed up at any of the 73 public primary care clinics under the Hospital Authority of Hong Kong (HAHK) were reviewed. Evidence-based audit criteria and performance standards were established after thorough literature review. In the first phase from 1^st^ April 2017 to 31^st^ March 2018, deficiencies of care were identified. It was followed by a one-year implementation phase through which a series of improvement strategies were executed. Outcome of the service enhancement was assessed in the second phase from 1^st^ April 2019 to 31^st^ March 2020. Student’s t test and Chi-square test were used to identify any statistically significant changes between the two.

**Results:** Totally 10,385 COPD cases were identified in Phase 1, among whom 3,102 (29.9%) were active smokers. Most of the patients were male (87.7%) and the mean age was 75.3±9.9 years old. Of those smokers, 1,788 (57.6%) had been referred to Smoking Counselling and Cessation Service (SCCS) and 1,578 (50.9%) actually attended it. 4,866 cases (46.9%) received Seasonal Influenza Vaccine (SIV) and 4,227 cases (40.7%) received Pneumococcal Vaccine (PCV). 1,983 patients (19.1%) had spirometry done before and 1,327 patients (12.8%) had been admitted to hospital due to acute exacerbation of COPD (AECOPD). With the concerted effort taken during the implementation phase, Phase 2 data showed a significant improvement in all criteria. There was a marked increase in the SIV and PCV uptake rate, spirometry performance rate and most importantly, a significant reduction in AECOPD rate leading to hospital admission (9.6%, P<0.00001).

**Conclusion:** COPD care at all public primary care clinics of HAHK had been significantly improved for all audit criteria via the systematic team approach, which in turn reduced the hospital admission rate and helped relieve the burden of the healthcare system.

## Background

Chronic obstructive pulmonary disease (COPD) is a progressive lung disease characterized by a persistent reduction of airflow resulting from chronic inflammation in the lung and remodeling of small airways (1). According to The Global Burden of Disease Study, there were 251 million COPD cases in 2016 around the world and around 3.2 million people died from COPD in 2015 (2). In Hong Kong (HK), the Population Health Survey 2014/15 reported that 0.5% of non-institutionalized persons aged 15 and above had doctor-diagnosed COPD (3). The disease accounted for over 30,000 episodes of inpatient discharges and deaths in 2016 and 1,223 registered deaths in 2017 (4). Therefore, COPD imposed a substantial economic and social burden to the health care system.

COPD is a commonly encountered condition in primary care. Primary care professionals have an essential role in taking comprehensive measures to improve the disease control and prevent its acute exacerbation. For example, large amount of evidence in the literature have shown that smoking cessation is the most effective intervention to slow down the disease progression of COPD (5). In addition, early diagnosis with spirometry test and implementation of influenza and pneumococcal vaccination have all been shown to reduce the disease burden and improve COPD patients’ quality of life (6-8). Despite all these evidences, the management of COPD is still far from satisfactory. For example, Kester et al. found that only 5% of Canadian general practitioners requested a pulmonary function test when attending an individual with clear signs of COPD (9). It is also disappointing to note that the referral rate and attendance rate for smoking cessation counselling service among COPD smokers remained low (10,11) and therefore their smoking cessation rate had been much poorer than those without COPD. Furthermore, the low take-up rate of seasonal influenza vaccine at 20–60% (12-15) will render this vulnerable group of patients a higher risk of developing chest infection during the winter surge. If all these preventive measures are not effectively implemented, acute exacerbation of COPD (AECOPD) would be inevitable. Indeed, acute exacerbations have been proven to lead to accelerated decline in lung function, increased COPD-related hospitalizations and mortality, and lastly augmented the health care utilization (16).

In HK, about 80% of COPD patients are managed under specialist care and 20% are managed at primary care clinics under the Hospital Authority of Hong Kong (HAHK) (17). A pilot survey conducted in 2011 found that COPD care at both primary and secondary level in HAHK needed to be improved (18). For example, spirometry was underused in the diagnosis and monitoring of patients with COPD (19) and a suboptimal adherence to accredited COPD management guideline was identified at five tertiary respiratory centers from 2013 to 2015 (20). Insufficient patient education, suboptimal coding of lung function results, underutilization of long-acting bronchodilators and lack of an integrative management model were the main issues to be tackled (21). In view of this, starting from 1 April, 2017, COPD audit had been conducted across all primary care clinics of HAHK to review the performance of COPD care so as to improve its clinical outcome. This study aimed to audit the management of COPD cases from all primary care clinics of HAHK and to work out improvement strategies. We believe that by improving the standard of care to COPD patients managed in the community via the aligned audit approach, the disease burden including the number of hospital admissions due to AECOPD would be greatly reduced.

## Method

### Study design

A two-phase clinic audit conducted at all 73 public primary care clinics of the HAHK.

### Setting audit criteria and justification of audit standards

The Quality Assurance (QA) subcommittee is a functional subcommittee under the leadership of Coordination Committee of Family Medicine [COC (FM)] in HAHK. Its main mission is to promote evidence based practice and to enhance the quality of care for all patients managed at General Outpatient Clinics (GOPCs) of HAHK. In late 2016, the QA subcommittee of COC (FM) agreed to conduct COPD audit across all GOPCs to improve the COPD care. Guidelines on COPD management published in recent 3 years were identified from the PubMed (https://pubmed.ncbi.nlm.nih.gov/). Members of QA Subcommittee, who were specialist family physicians, designed six evidence-based audit criteria for COPD care after through literature review (22-23). The audit criteria and performance standards are summarized in **Table 1**.

**Table 1.** list of audit criteria and performance standards of the study.

| Item no. | Recommendations and audit criteria | Standard |
| --- | --- | --- |
| 1 | Recommendations: All COPD patients should be regularly reviewed with a pre-scheduled appointment in General Outpatient Clinic (GOPC);<br>Criteria 1: % of COPD patients with pre-scheduled appointments in GOPC. | 85% |
| 2 | Recommendations: All smoker COPD patients should be advised on smoking cessation and referred to Smoking Counselling and Cessation Services (SCCS);<br>Criteria 2: COPD patients who are smokers:<br>a. % of COPD patients referred to SCCS before or<br>b. % of COPD patients ever attended SCCS before | 55%<br>50% |
| 3 | Recommendations: All COPD patients should receive seasonal influenza vaccine (SIV) annually unless contraindicated;<br>Criteria 3: % of COPD patients who have received SIV in the preceding year. | 50% |
| 4 | Recommendations: All COPD patients should receive pneumococcal vaccine (PCV) unless contraindicated;<br>Criteria 4: % of COPD patients who have received PCV before. | 50% |
| 5 | Recommendations: Spirometry is recommended for all COPD cases at diagnosis or when the alternative diagnosis needs to be ruled out, and to monitor the disease progression.<br>Criteria 5: % of COPD patients with spirometry test done before. | 50% |
| 6 | Recommendations: All efforts should be made to reduce the acute exacerbation of COPD (AECOPD).<br>Criteria 6: % of COPD patients admitted to hospitals due to AECOPD. | <10% |

### Data collection and analysis

#### Inclusion criteria

Eligible study subjects were COPD patients aged 40 or above and had attended any of the 73 GOPCs of HAHK for follow-up (FU) during the study period. Locally, the HAHK has been responsible for managing HK’s public hospitals since 1991. There are 7 Family Medicine Departments in 7 hospital clusters of HAHK, and totally 73 GOPCs are under its jurisdiction. The majority of patients attending the GOPCs for medical care are the elderly, the low income group and those with chronic diseases. The diagnosis of COPD was identified through the computerized patient record, the Clinical Management System (CMS), of the HAHK. Relevant consultations were identified by the International Classification of Primary Care (Second edition) code of ‘R79-Chronic bronchitis’ or ‘R95-Chronic obstructive pulmonary disease’ over a 12-month reporting period. COPD patients managed in the hospital Specialist Outpatient Clinics (SOPCs) or certified dead during the study period were excluded from the data analysis.

#### The audit cycle

Phase 1 was from 1^st^ April 2017 to 31^st^ March 2018, with deficiencies of care identified. To fill in the service gaps, one-year implementation phase was carried out from 1^st^ April 2018 to 31^st^ March 2019, through which a series of improvement strategies were executed. Outcome of the enhancement was reviewed during Phase 2 from 1^st^ April 2019 to 31^st^ March 2020.

##### First-phase data collection and analysis

A COPD patient registry had been retrieved from the Clinical Data Analysis and Reporting System (CDARS) of HAHK by Head Office Statistical Team. Total 12,003 COPD patients were identified during Phase 1. Among them, 454 cases were certified dead (annual all-cause mortality rate 3.8%) and 1,164 cases were found to have FU at SOPCs and therefore were excluded. The remaining 10,385 cases (86.5%) fulfilling the inclusion criteria were included into data analysis.

##### Implementing changes and intervention

The QA subcommittee of COC (FM) is composed of department head and representatives from each Family Medicine Department in the 7 clusters of HAHK, all of whom are specialist family physicians. COPD audit working group led by the QA subcommittee member of each cluster was formed on 1^st^ April 2017 and subsequently, doctor and nurse subject officers were assigned. A structured team approach was adopted to set up strategies on administrative policy and management guidelines. The COPD patient statistics were shared among clinic doctors and nurses at least annually. Individual clinic staffs then work out their local FU actions. Regular service review meeting had been conducted quarterly to half yearly.

At the practice level, a series of computer-based enhancement in the CMS was initiated to facilitate the COPD data capture. For example, a specifically designed “COPD module” was launched out in the CMS of all GOPCs in 2016. In this module, all COPD related data including the epidemiology data, spirometry test results and disease severity groupings etc. were recorded. Moreover, CMS reminder had been set up in almost all GOPCs to enhance the SCCS referral and vaccination upon patient’s registration. Apart from the CMS enhancement, the workflow of COPD care in some GOPCs had also been revamped to meet the audit standard. For example, COPD cases would be referred to attend a nurse-led assessment upon their routine FU, where a comprehensive range of services would be provided including nursing counselling, smoking cessation service referral if smoker, and a course of pulmonary rehabilitation by allied health workers including physiotherapist and occupational therapist. In addition, all doctors were recommended to refer COPD cases for spirometry test at diagnosis and to monitor the disease progression. More office spirometry machines had been purchased in some clusters to meet the increasing service demand on spirometry assessment. COPD cases would then receive standard assessment including the Modified Medical Research Council (mMRC) dyspnea scale and COPD Assessment Test (CAT) score evaluation, and were graded according to the definition of GOLD guideline (22).

At the clinic level, a policy on COPD risk factor screening was advocated, and a continuous monitoring-and-feedback system with ongoing problem solving was reinforced. Regular review on the progress of the audit was carried out where feedback regarding the deficiencies was tackled promptly. For example, the performance of each GOPC on COPD care would be reviewed at least biannually with loopholes identified. Meanwhile, the good practice from other GOPCs will be shared out for cross learning purpose. At the doctor level, diagnosis and management of COPD and prevention of acute exacerbation based on the latest GOLD guideline had been promulgated to all frontline doctors to sharpen their skill set. All doctors were advised to manage COPD cases according to their severity as suggested by the grading system, and to make appropriate referrals if deemed necessary. At patient level, regular health talk had been organized by various ranks of staff to empower the patients on self-care and they were advised to seek medical care immediately should symptoms suggestive of AECOPD occur. All COPD smoker patients would be advised to receive the smoking cessation counselling service unless refusal to participate. The deficiencies in service provision and corresponding implementation strategies are summarized in **Table 2**.

**Table 2.**
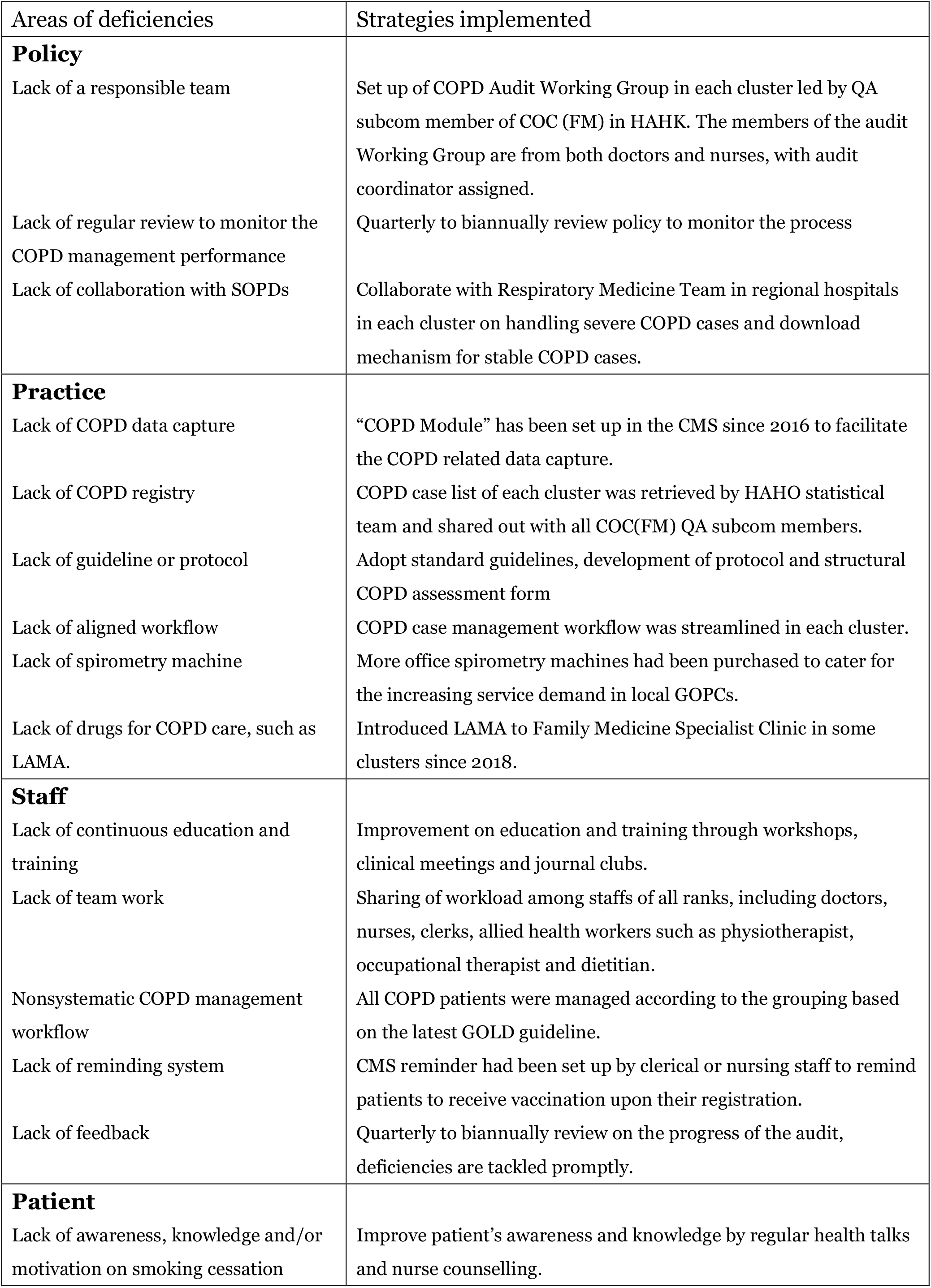
Summaries of deficiencies identified and strategies implemented.

| Areas of deficiencies | Strategies implemented |
| --- | --- |
| **Policy**<br>Lack of a responsible team<br>Lack of regular review to monitor the COPD management performance<br>Lack of collaboration with SOPDs | Set up of COPD Audit Working Group in each cluster led by QA subcom member of COC (FM) in HAHK. The members of the audit Working Group are from both doctors and nurses, with audit coordinator assigned.<br>Quarterly to biannually review policy to monitor the process<br>Collaborate with Respiratory Medicine Team in regional hospitals in each cluster on handling severe COPD cases and download mechanism for stable COPD cases. |
| **Practice**<br>Lack of COPD data capture<br>Lack of COPD registry<br>Lack of guideline or protocol<br>Lack of aligned workflow<br>Lack of spirometry machine<br>Lack of drugs for COPD care, such as LAMA. | “COPD Module” has been set up in the CMS since 2016 to facilitate the COPD related data capture.<br>COPD case list of each cluster was retrieved by HAHO statistical team and shared out with all COC(FM) QA subcom members.<br>Adopt standard guidelines, development of protocol and structural COPD assessment form<br>COPD case management workflow was streamlined in each cluster.<br>More office spirometry machines had been purchased to cater for the increasing service demand in local GOPCs.<br>Introduced LAMA to Family Medicine Specialist Clinic in some clusters since 2018. |
| **Staff**<br>Lack of continuous education and training<br>Lack of team work<br>Nonsystematic COPD management workflow<br>Lack of reminding system<br>Lack of feedback | Improvement on education and training through workshops, clinical meetings and journal clubs.<br>Sharing of workload among staffs of all ranks, including doctors, nurses, clerks, allied health workers such as physiotherapist, occupational therapist and dietitian.<br>All COPD patients were managed according to the grouping based on the latest GOLD guideline.<br>CMS reminder had been set up by clerical or nursing staff to remind patients to receive vaccination upon their registration.<br>Quarterly to biannually review on the progress of the audit, deficiencies are tackled promptly. |
| **Patient**<br>Lack of awareness, knowledge and/or motivation on smoking cessation | Improve patient’s awareness and knowledge by regular health talks and nurse counselling. |

##### Second-phase data collection and analysis

A total of 11,592 COPD patients were found to have regular FU in GOPCs of HA during Phase 2. Of those being excluded in this phase, 404 cases were certified dead (annual all-cause mortality rate 3.5%) and 1,675 cases were found to have FU at SOPDs. The remaining 9,513 cases (82.1 %) were included into data analysis.

#### Determination of variables

The recruited patients’ age, gender and smoking status were retrieved from the CMS of the HAHK. All data of the 6 audit criteria were retrieved from the CDARS of HAHK by head office statistics team. An appointment is said to be pre-scheduled if the date of booking is at least 3 calendar days earlier than the date of appointment. A smoker is said to be referred to SCCS if he/she had active appointment booked for SCCS. SIV and PCV vaccination rate were retrieved from the CMS immunization module. Criteria 6 on COPD patients admitted to hospitals due to AECOPD refers to the discharged episodes with principal diagnosis of COPD exacerbation during the reporting period. The ICD-9 diagnosis codes for COPD exacerbations leading to hospital admission are listed in the **appendix**.

### Statistical methods

All data were entered and analyzed using computer software (SPSS version 16.0; Inc., Chicago [IL], US). Chi-square test and independent student’s t test were used respectively for categorical and continuous variables to examine the statistically significant differences between the first phase and second phase measurements. A P value of <0.05 (two-tailed) was regarded as statistically significant.

## Results

**Table 3** summarizes the demographic characteristics of COPD patients included into the two phases. Their age, gender and smoking status were comparable. Around 88% of the COPD patients in both phases were male and the majority were elderly patients aged over 65 years old. Approximately 30% of COPD patient were chronic smokers.

**Table 3.** Demographic characteristics of COPD patients in the two phases.

|  | Phase 1 | Phase 2 | P value |
| --- | --- | --- | --- |
| COPD cases included in data analysis (n) | 10,385 | 9,513 | / |
| Gender |  |  | 0.53 |
| Male | 9,108 (87.7%) | 8,371 (88.0%) |  |
| Female | 1,277 (12.3%) | 1,142 (12.0%) |  |
| Age (years) | 75.3 ± 9.9 | 75.1 ± 9.8 | 0.92 |
| <40-49 | 117 (1.1%) | 92 (1.0%) | 0.12 |
| 50-64 | 1,559 (15.0%) | 1,450 (15.2%) |  |
| 65-79 | 5,031 (48.4%) | 4,732 (49.7%) |  |
| ≥80 | 3,678 (35.4%) | 3,239 (34.0%) |  |
| Smoker | 3,102 (29.9%) | 3,008 (31.6 %) | 0.08 |

Data are shown as No. (%) of cases or mean ± standard deviation.

Among the 10,385 patients reviewed in Phase 1, 1,144 cases (11.0%) were referred to specialists for continued care. Among the 9,153 patients included in Phase 2, 6,088 patients were FU cases from Phase 1, with a case overlapping rate of 64.0%. At the end of the Phase 2, 1,057 cases (11.1%) were referred to specialists for further care. The SOPD referral rate was also comparable between the two phases (P=0.83).

A comparison of the standards achieved in the two phases is summarized in **Table 4**. In the first phase, marked deficiencies were identified in almost all criteria. It is quite alarming to note that less than one fifth of COPD patients (19.1%) had performed spirometry before. 1,327 cases (12.8%) had at least one episode of hospital admission due to AECOPD. After proactive execution of the enhancement strategies during the implementation phase, significant improvement was observed with respect to all criteria in Phase 2. The most clinically important change was observed in the hospital admission rate due to AECOPD, reducing to 910 (9.6%) in Phase 2 (P<0.00001). Further analysis on the frequencies of hospital admissions due to AECOPD revealed that the reduction had been significant for all frequencies including being admitted once, twice, and three times or more between the two phases (P<0.00001).

**Table 4.**
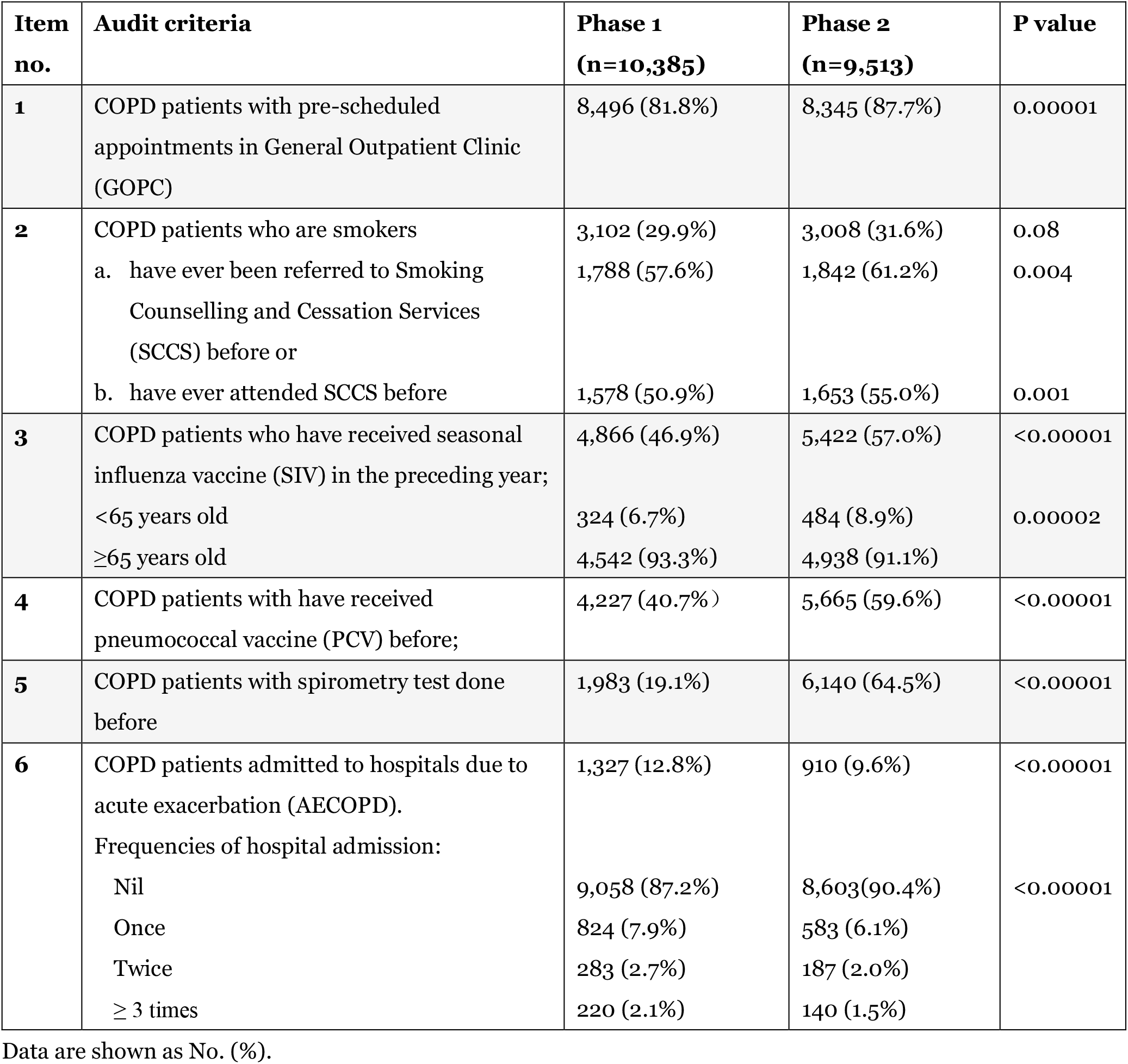
Number and percentage of patients with criteria fulfilled in Phase 1 and Phase 2 and comparison of the results in the two phases.

| Item no. | Audit criteria | Phase 1<br>(n=10,385) | Phase 2<br>(n=9,513) | P value |
| --- | --- | --- | --- | --- |
| 1 | COPD patients with pre-scheduled appointments in General Outpatient Clinic (GOPC) | 8,496 (81.8%) | 8,345 (87.7%) | 0.00001 |
| 2 | COPD patients who are smokers | 3,102 (29.9%) | 3,008 (31.6%) | 0.08 |
|  | a. have ever been referred to Smoking Counselling and Cessation Services (SCCS) before or | 1,788 (57.6%) | 1,842 (61.2%) | 0.004 |
|  | b. have ever attended SCCS before | 1,578 (50.9%) | 1,653 (55.0%) | 0.001 |
| 3 | COPD patients who have received seasonal influenza vaccine (SIV) in the preceding year; | 4,866 (46.9%) | 5,422 (57.0%) | <0.00001 |
|  | <65 years old | 324 (6.7%) | 484 (8.9%) | 0.00002 |
|  | ≥65 years old | 4,542 (93.3%) | 4,938 (91.1%) |  |
| 4 | COPD patients with have received pneumococcal vaccine (PCV) before; | 4,227 (40.7%) | 5,665 (59.6%) | <0.00001 |
| 5 | COPD patients with spirometry test done before | 1,983 (19.1%) | 6,140 (64.5%) | <0.00001 |
| 6 | COPD patients admitted to hospitals due to acute exacerbation (AECOPD). | 1,327 (12.8%) | 910 (9.6%) | <0.00001 |
|  | Frequencies of hospital admission: |  |  |  |
|  | Nil | 9,058 (87.2%) | 8,603(90.4%) | <0.00001 |
|  | Once | 824 (7.9%) | 583 (6.1%) |  |
|  | Twice | 283 (2.7%) | 187 (2.0%) |  |
|  | ≥ 3 times | 220 (2.1%) | 140 (1.5%) |  |

Data are shown as No. (%).

Further comparisons on the hospital admission rate due to AECOPD revealed that the reduction was only significant among the elderly age groups (from 14.1% in phase 1 to 10.5% in phase 2, P<0.00001), whereas the improvement in the younger age group was not significant (P=0.2) (**Figure 1**).

**Figure 1.**
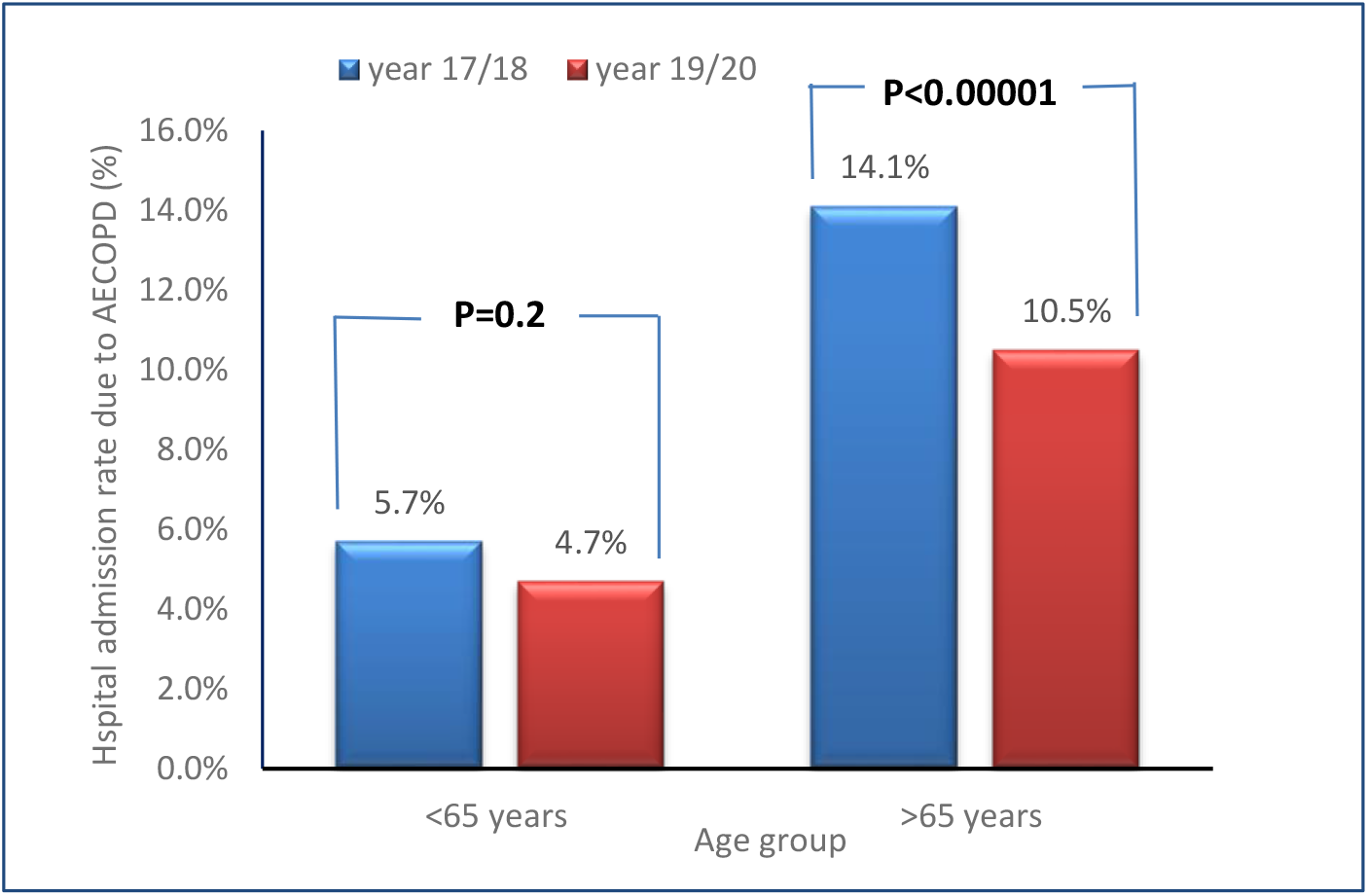
Comparison of hospital admission rate due to AECOPD among different age groups between the two phases.

## Discussion

Marked deficiencies existed in various aspects of COPD management in the primary care setting in phase 1. This is an expected albeit disappointing finding due to the service gaps at different levels such as knowledge set, skill set and tool set. These were particularly apparent in the process criteria such as smoking cessation service referral, uptake rate of SIV or PCV, and under-utilization of spirometry test. Through the concerted effort paid during the audit process, COPD management at all public primary care clinics in HK had been greatly enhanced, with all audit criteria being tremendously improved and target audit standard achieved. The outcome criteria, AECOPD rate leading to hospital admission, was also significantly reduced in Phase 2.

It is of note that the overall headcounts of COPD patients under the care of HAHK had been decreasing over the years. In view of the comparable annual overall mortality rate (3.8% in phase 1 versus 3.5% in phase 2, P=0.22) and SOPD referral rate (11.0% in phase 1 versus 11.1% in phase 2, P=0.83) between the two phases, we believe that the reduction in the total case number of COPD was likely due to the decreased prevalence of COPD in HK although the exact data is still lacking. Indeed, an overview on the prevalence and incidence of chronic respiratory disease published early this year showed that the incidence rate of COPD decreased in all age groups from 1990 to 2017 globally, except for elderly patients older than 80 years (24). This reduced prevalence could partially be explained by the fact that the prevalence of daily cigarette smokers of HK dropped to 10.0% in 2017, which is the lowest rate recorded since 1982 according to the Thematic Household Survey released by Census and Statistics Department of HK (25).

About 20% of COPD patients were found not to have a prescheduled FU for regular assessment at the beginning of the audit. Currently there is no cure for COPD, but treatment can help control the symptoms and slowdown its progression, therefore warranting a regular FU to assess its severity. On the one hand, all doctors were reminded to arrange an FU appointment for all COPD cases during their routine consultation. On the other, the audit team had gone through the COPD case registry who had no FU appointment in the CMS to review whether they had been arranged to have any FUs with private doctors or respiratory specialists. Some were called up individually by nursing staff to enquire about their symptom control. A FU appointment would be offered if the patient had not received proper assessment in the recent one year. With such effort, many lost-to-FU COPD cases returned to GOPCs for spirometry and clinical assessment, with 87.7% of all COPD cases having regular FU in GOPCs in Phase 2 (P<0.00001).

Smoking leads to COPD in more than 80% of cases and smoking cessation is the most effective intervention in stopping the progression of COPD, as well as increasing the survival and reducing the morbidity (26). In view of this, evidence based approach using a combination of behavioral and pharmaceutical interventions had been adopted at the SCCS in HAHK since 2010. As a routine practice, all primary care doctors refer COPD smokers to the SCCS during the consultation. However, evidence-practice gap did exist and it is not surprising to find that only about half of COPD smokers were referred and even fewer actually attended the SCCS in Phase 1. These findings were consistent with literature suggesting that COPD smokers are poorly motivated to quit smoking in general (10). Another possibility is the physician-related factor. For example, literature shows that some physicians do not routinely deal with smoking cessation during their consultations with smokers partly due to lack of cessation specific knowledge or skills and partly due to insufficient consultation time (27). This is particularly true in HK where the average time allocated for each consultation is only about 7 minutes at GOPCs. To address this service gap, a series of training programs on smoking cessation counseling and motivational interview had been organized to strengthen staff’s knowledge and skill on smoking cessation. Besides, the SCCS workflow had been revamped in some clusters so that the counselling could be delivered in the same session once referred, which resulted in a significantly reduced default rate. With the concerted effort and cohesive support from the smoking cessation nurse, the situation at Phase 2 had been significantly improved with the target standard achieved.

For criteria 3 and 4 on the update rate of SIV and PCV, although our performance had significantly improved during the audit cycle, only more than half of COPD patients were vaccinated against SIV (57.0%%) and PCV (59.6%) in Phase 2. Indeed, influenza vaccination coverage rates among COPD patients remain low in many countries (13-14). In HK, all elderly patients aged over 65 years are entitled to receive SIV and PCV for free under the HK Government Vaccination Program. However, the breakdown figures of SIV coverage among those under age 65 were still far from satisfactory (6.7% in Phase 1 and 8.9% in Phase 2). Given the widely established evidence on the long-term benefits of SIV on COPD care, such as reduced number of exacerbations, hospitalizations and all-cause mortalities (7), we would like to propose that HK government should launch out free SIV to COPD patients of all ages to reduce the mortality.

It is alarming to find that only 19.1% COPD patients had spirometry test before in Phase 1. This was consistent with local studies done in 2013 showing that spirometry had been underused in general but especially by family physicians in the management of COPD patients, with only 18.3% patient had spirometry done during diagnostic workup (19).

The reasons accounting for this poor performance rate are multifactorial. At doctors’ level, some doctors often make the diagnosis of COPD based on clinical features alone. At clinic level, spirometry service was previously only available at hospital setting. Therefore, suspected COPD patients had to be referred to Respiratory Specialist Clinic to perform the lung function tests where the waiting time ranged from months to two years under HAHK. To plug this loophole, a series of education talks on the proper diagnosis and management of COPD, emphasizing on the importance of spirometry test, were delivered to all doctors. Furthermore, some selected GOPCs were equipped with spirometry machine during the implementation phase so that the spirometry test could be conveniently performed locally within 2-4 weeks. In addition, at least 1-2 designated nurses from these GOPCs had been specially trained on how to perform the spirometry correctly based on the aligned standard. With such facilitations both on the skill set and tool set, tremendous improvement was observed for this criteria in Phase 2 (64.5%, P<0.00001).

The last criteria 6, the rate of AECOPD leading to hospital admission, is the single most important outcome criteria of this audit. Mild to moderate AECOPD that had been well managed in GOPCs would not be included in this criteria. In Phase 1, almost 1 in 8 of COPD patient (12.8%) had been admitted to hospital due to AECOPD. This rate was lower than those reported in the literature which showed around 20% of COPD patients had experienced severe acute exacerbations leading to hospital admission annually (28, 29). This discrepancy might be explained by the fact that majority of COPD cases managed in primary care clinics belong to the mild grade (Group A and B) according to GOLD guideline. Indeed, local data from Kowloon Center Cluster of HAHK has confirmed that only about 10% of COPD cases FU in primary care clinics belonged to Group C and D (30). In order to further decrease the burden of hospital admissions, prevention and prompt treatment of exacerbations are the key goals in COPD care. With such mindset, a series of service enhancement strategies were executed. Firstly, early identification of COPD patients by spirometry and proper grading according to the GOLD guideline were done as mentioned above. Secondly, all COPD cases were managed according to their grading, hence providing the right level of care to the right patients. For example, stable Group A/B patients would continue regular FU at GOPCs, where long acting anti-muscarinic antagonist (LAMA) was newly introduced in 2018 in some clusters to improve the symptom control. For more severe Group C/D patients that warrant advanced care, a timely referral to the Respiratory Specialists would be initiated. Lastly, relatively stable AECOPD patients had been managed locally with more frequent FU instead of being admitted to hospital, hence reducing the hospital burden. This is in line with findings in the literature that frequent outpatient visits prevent the exacerbation of COPD (31). With all these proactive interventions and efforts, the AECOPD rate leading to hospital admission was significantly reduced to 9.6% in Phase 2. In addition, the reduction was significant for COPD patients admitted to hospital at all frequencies, proving that our strategies were effective for COPD patients of different severities.

### Strength and Limitations of this study

COPD patients are one of the most vulnerable group of population in the community. COPD, by its disease nature, constitutes a substantial burden to the health care system of all countries in the world. Therefore, clinical audit on this important topic and the subsequent continuous quality improvement programs will likely to have tremendous impact on COPD care in the community. To our knowledge, this study is one of the biggest clinical audit on COPD management ever conducted internationally, and it has provided crucial information on COPD care in primary care setting. Its wide coverage (all 73 public primary care clinics in HAHK) and large sample size (around 10,000 cases in both Phase 1 and 2) have significantly increased the statistical power of the study. The broad spectrum of audit criteria evaluated in the study reflected the comprehensive nature of COPD management in primary care. In addition, all audit criteria were based on objective assessment parameters with data being retrieved from the computer system of HAHK, therefore minimizing the confounding effect due to recall basis or data entry error.

With that being said, this study has several limitations. Firstly, the study was carried out in public primary care clinics of HAHK, therefore the results may not be applicable to the private sector or secondary care setting. Nevertheless, since COPD cases from all GOPCs of HAHK had participated in the clinic audit, these data should realistically represent the COPD care in public primary care settings and provided important background information for future service enhancement. Secondly, this clinical audit mainly focused on short-term outcome aspects of COPD management. Long-term outcomes such as smoking cessation rate, lung function improvement or COPD-related mortality rate were not analyzed. In addition, process indicator such as assessment on inhaler technique was not included. This gap will need to be filled as evidence has revealed that inhaler technique education significantly reduced the acute exacerbation rate (32). Subsequent studies focusing on the long-term outcome criteria and inhaler technique may help provide a more thorough picture of COPD management.

## Conclusion

COPD management at all primary care clinics of HAHK had been significantly enhanced during the past three years. Via a team approach with streamlined governance and structure as well as proactive staff engagement, marked improvement had been achieved in all audit criteria for COPD management. The significant reduction in AECOPD rate will help relieve the burden to specialist care and hospital in the long run.

## Data Availability

The datasets used in the current study were compiled by the Statistical Team from the Head Office of HAHK and are used as internal reference only. They would be available from the corresponding author on reasonable request after being approved by Head Office of HAHK.

## Acknowledgement

We would like to thank all clinical staff of 73 GOPCs under the seven clusters of the HAHK, namely the Dept. of Family Medicine and Primary Health Care of Hong Kong East Cluster, Hong Kong West Cluster, Kowloon East Cluster, Kowloon Central Cluster, Kowloon West Cluster, New Territories East Cluster and New Territories West Cluster, for their professional service and unfailing support to this clinic audit, without which the significant service enhancement would not have been achieved. We are also indebted to all members of COC (FM) QA Subcommittee for her leadership in setting up the audit criteria and members of Primary Health Care Team of the Head Office of HAHK for their tremendous support. Our gratitude also goes to Head Office Statistical Team of HAHK and Ms. Katherine CHAN, Statistical Officer I of Queen Elisabeth Hospital of Kowloon Central Cluster for their expertise in retrieving the data from the CMS and CDARS of HAHK.

## Abbreviations

AECOPD: Acute Exacerbation of Chronic Obstructive Pulmonary Decease
CAT: COPD Assessment Test
CDARS: Clinical Data Analysis and Reporting System
CMS: Clinical Management System
COPD: Chronic Obstructive Pulmonary Disease
FU: Follow Up
GOLD guideline: Global Initiative for Obstructive Lung Disease guideline
GOPCs: General Outpatient Clinics
HAHK: Hospital Authority of Hong Kong
HK: Hong Kong
LAMA: Long Acting Anti-muscarinic Antagonist
mMRC: Modified Medical Research Council
PCV: Pneumococcal Vaccine
QA subcommittee of COC (FM): Quality Assurance subcommittee of the Coordination Committee of Family Medicine
SCCS: Smoking Counselling and Cessation Service
SIV: Seasonal Influenza Vaccine
SOPCs: Specialist Outpatient Clinics

## Appendix List of ICD-9 Diagnosis Codes for COPD Exacerbations at hospital admission

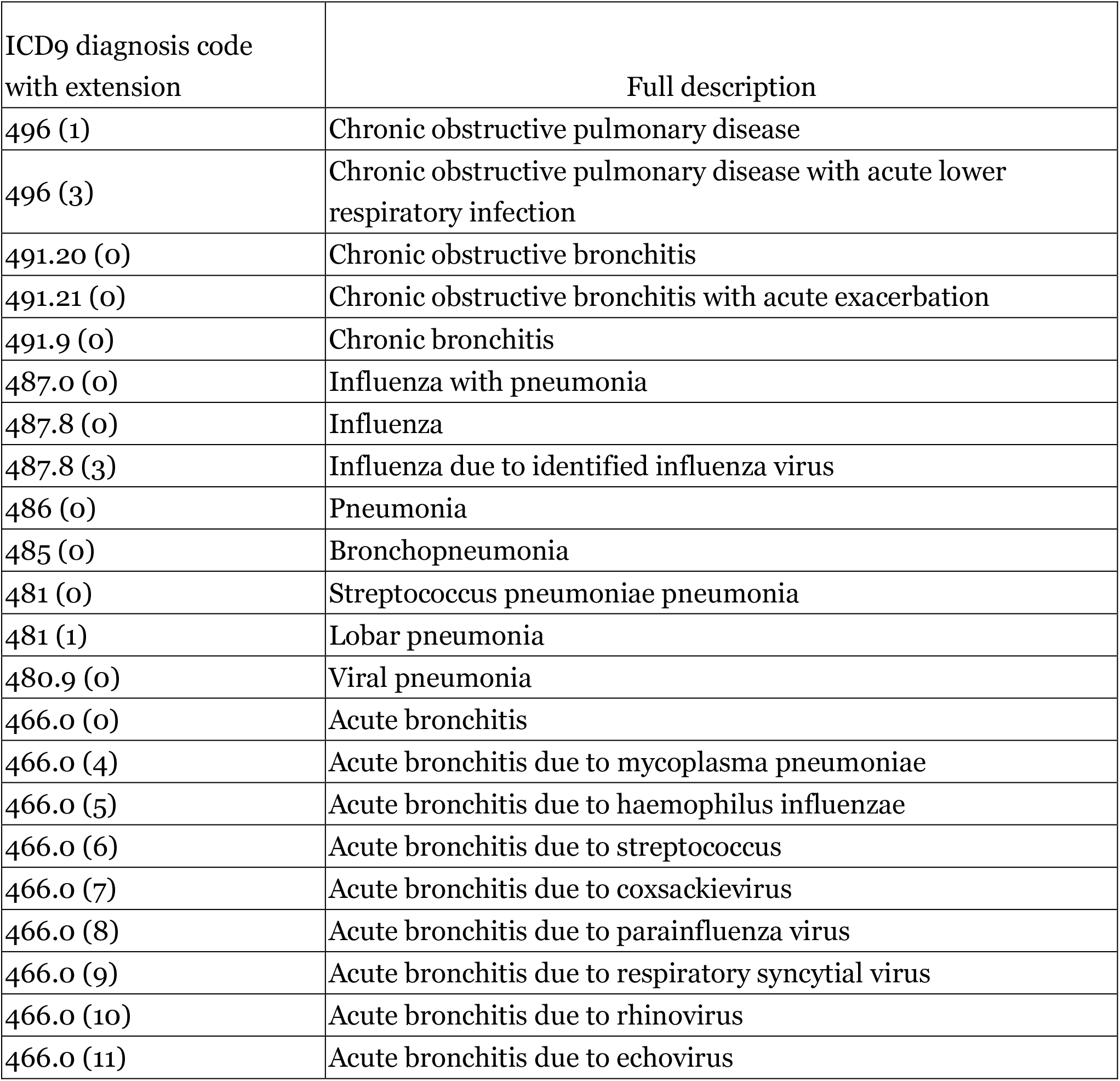

Appendix: List of ICD-9 Diagnosis Codes for COPD Exacerbations at hospital admission
| ICD9 diagnosis code with extension | Full description |
| --- | --- |
| 496 (1) | Chronic obstructive pulmonary disease |
| 496 (3) | Chronic obstructive pulmonary disease with acute lower respiratory infection |
| 491.20 (0) | Chronic obstructive bronchitis |
| 491.21 (0) | Chronic obstructive bronchitis with acute exacerbation |
| 491.9 (0) | Chronic bronchitis |
| 487.0 (0) | Influenza with pneumonia |
| 487.8 (0) | Influenza |
| 487.8 (3) | Influenza due to identified influenza virus |
| 486 (0) | Pneumonia |
| 485 (0) | Bronchopneumonia |
| 481 (0) | Streptococcus pneumoniae pneumonia |
| 481 (1) | Lobar pneumonia |
| 480.9 (0) | Viral pneumonia |
| 466.0 (0) | Acute bronchitis |
| 466.0 (4) | Acute bronchitis due to mycoplasma pneumoniae |
| 466.0 (5) | Acute bronchitis due to haemophilus influenzae |
| 466.0 (6) | Acute bronchitis due to streptococcus |
| 466.0 (7) | Acute bronchitis due to coxsackievirus |
| 466.0 (8) | Acute bronchitis due to parainfluenza virus |
| 466.0 (9) | Acute bronchitis due to respiratory syncytial virus |
| 466.0 (10) | Acute bronchitis due to rhinovirus |
| 466.0 (11) | Acute bronchitis due to echovirus |

## Notes

### Competing Interest Statement

The authors have declared no competing interest.

### Funding Statement

All authors have completed the Unified Competing Interest form and declare: no support from any organisation for the submitted work; no financial relationships with any organisations that might have an interest in the submitted work in the previous three years, no other relationships or activities that could appear to have influenced the submitted work. This project was not funded by any agency in the public, commercial, or not-for-profit sectors. All authors have declared no conflicts of interest.

### Author Declarations

No ethical approval is needed in view of audit nature of the study. This complies with local ethics committee from the Hospital Authority of Hong Kong that no formal ethical approval was required for clinical audit.

